# Individual, Household and Contextual Factors Associated with Anaemia in Children in Nigeria

**DOI:** 10.1101/2024.06.20.24308990

**Authors:** Tukur Dahiru, Suleiman Usman, Khadeejah Liman Hamza, Bilkisu Nwankwo, Awawu Grace Nmadu, Aminu Lawal, Idris Muhammad Yakubu, Adegboyega Moses Oyefabi

**Affiliations:** Ahmadu Bello University Zaria, Kaduna State, Nigeria; Ahmadu Bello University Teaching Hospital, Shika-Zaria, Kaduna State, Nigeria; Kaduna State University, Kaduna, Kaduna State, Nigeria; Katsina State Primary Health Care Agency Katsina, Katsina State, Nigeria; CBN Diagnostic and Treatment Center Bauchi, Bauchi, Nigeria

**Keywords:** Anaemia, Child, Deficiency, Haemoglobin, Health, Iron, Malnutrition, Micronutrient, Public

## Abstract

**Introduction:** Anaemia among children is an issue of significant concern within the realm of public health due to its potential to result in significant adverse outcomes, including cognitive deficits and growth deficiencies. Its prevalence exhibits regional variations, wherein larger frequencies are observed in regions characterized by lower levels of socioeconomic development. This study sought to identify the individual, household, and contextual factors associated with anaemia in Nigeria.

**Methods:** Data on a weighted sample of 11,223 children under five years of age from 42,000 households was obtained from the Nigeria Demographic and Health Survey (NDHS) 2018. The study utilized the household members recode file, a data base which allows for the estimation of prevalence of childhood anaemia together with selected household variables and the kids recode file, a data base that allows for estimation of other household variables. Relevant variables were then generated, and the two data files were merged. Descriptive statistics were generated, and the multi-level logistic regression was computed to identify individual, household and contextual factors associated with anaemia.

**Results:** The study found that the overall prevalence of anaemia was 67.9%. It was noted that being a: child of Muslim parents; big baby at birth; wasted, stunted; underweight, child resident in Southeast and Southwest Nigeria and a child resident in rural area were significantly associated with increased odds of anaemia. While having a mother with secondary education, being a child in the second, third, or fourth birth order belonging to richest households, living in households with tiled floors, living in households with cemented floors, living in households with carpeted floors and improved sanitation/water source and living in Northeast Nigeria were significantly associated with decreased odds of anaemia. About of 8.5% individual, household and contextual factors determine the average prevalence of anaemia in children in Nigeria.

**Conclusion:** The findings from this study reveal that individual, household and contextual factors are important determinants of anaemia in children in Nigeria.

## Introduction

The prevalence of anaemia among children is an issue of significant concern within the realm of public health due to its potential to result in significant adverse outcomes, including cognitive deficits and growth deficiencies. For instance, anaemia resulting from iron deficiency can hinder the process of linear growth and contribute to stunting.^1^ The significant association between iron deficiency anaemia and the impairment of cognitive development in children has been linked with poor academic performance, potentially impacting the child’s future opportunities.

According to the World Health Organization (WHO), anaemia in children is characterized by low levels of haemoglobin (<11g/dl) and may manifest in different levels of severity, ranging from mild to severe.^2^ The most prevalent global causes of this condition, as indicated by the 2019 Global Burden of Disease study, include dietary iron deficiency, vitamin A deficiency, and beta-thalassemia trait.^2^ The public health importance of anaemia is classified based on its prevalence into four categories: normal (≤4.9%), mild (5.0 – 19.9%), moderate (20.0 – 39.9%), and severe (≥40.0%). Its prevalence exhibits regional variations, wherein larger frequencies are observed in regions characterized by lower levels of socioeconomic development.^3^ Conversely, lower frequencies are observed in regions characterized by higher levels of socioeconomic development.

Findings of a systematic review of the prevalence of anaemia from 204 countries, showed that anaemia has a higher impact on children under the age of five, with a prevalence of 39.7% (95% CI: 39.0–40.4), in comparison to the prevalence of 22.8% (95% CI: 22.6–23.1) observed across all age groups.^3^ Within the sub-Saharan sub region, anaemia has persistently surpassed the worldwide average, with an overall incidence of 59.9% and an alarming 72.3%, specifically in Ethiopia.^4,5^ According to the findings of the 2018 Nigeria Demographic and Health Survey (NDHS), a significant proportion (68%) of children between the ages of 6 and 59 months, were affected by anaemia.^6^ This condition was further classified into three categories: mild anaemia, affecting 27% of the children; moderate anaemia, affecting 38% of the children; and severe anaemia, affecting 3%.^6^ In 2019, anaemia was responsible for 58.6 million years lived with disability (YLD) (95% CI: 40.14–81.1).^3^ The areas of Western Sub-Saharan Africa, South Asia, and Central Sub-Saharan Africa saw the greatest burdens with this condition.^3^ The statistics highlight the gravity of anaemia in the sub-Saharan African region.

Studies have also documented significant social and demographic characteristics associated with an increased likelihood of developing anaemia in sub-Saharan African countries.^4,7,8^ In a national representative survey in Tanzania factors associated with iron deficiency anaemia in children included maternal work status, level of education, Body Mass Index(BMI), anaemia status, place of delivery, sex of child, size of household, illness and child anthropometry.^1^ The NDHS of 2018 had highlighted some factors associated with anaemia in revealing higher prevalence among male children (69.5%), those with mothers in the lowest wealth quintile (80.1%), lacking formal education (75.1%), and residing in rural areas. It also unveils regional disparities in anaemia prevalence among children aged 6-59 months.^6^

Understanding the extent to which these factors contribute to development of anaemia is crucial for devising effective interventions for children.^4^ Though a few studies have examined factors associated with anaemia among children aged 6–59 months in Nigeria, there are limited studies that have explored the entire range of individual, household and contextual factors associated with in children aged 6-59 months^8^ For example, a study evaluating the predictors of anaemia among children aged 6–59 months in Nigeria did not encompass the impact of household feeding practices on anaemia occurrence^8^ Additionally, though the NDHS 2018 highlights the presence of some individual, household, and contextual factors associated with the prevalence of anaemia in children less than 6-59 months, these associations were not statistically validated using advanced techniques in the NDHS 2018 report, as observed in other studies.^1,4,5^ This study seeks to employ nationally representative data to identify the factors associated with anaemia at individual, household, and contextual levels among children in Nigeria.

## Materials and Methods

### Study design, Data sources, Study sample

We drew data from 2018 Nigeria Demographic and Healthy Survey (2018 NDHS) which was a cross-sectional study and produced a nationally representative sample of women of reproductive age group. The 2018 NDHS had a single objective of providing up-to-date basic demographic and health indicators among adult and adolescent populations as well as among children under-five years of age including prevalence of anaemia among this population. The 2018 NDHS was based on a two-stage cluster sampling where samples were selected independently in every stratum. Sampling was based on the 2006 Population and Housing Census of the Federal Republic of Nigeria (NPHC) enumeration areas (EAs), and these represented the primary sampling units (PSU). The PSU were used as the clusters for the 2018 NDHS and their list served as the sampling frame during field work. In the first stage, 1,400 EAs were selected with probability proportional to EA size. In the second stage’s selection, a fixed number of 30 households were selected in every cluster through equal probability systematic sampling, resulting in a total sample size of approximately 42,000 households. All women aged 15-49years in the selected households “who were either permanent residents of the selected households or visitors who stayed in the households the night before the survey” were interviewed.

A sub-sample of one-third of the sampled household was taken and men aged 15-59 years in these households were interviewed. Alongside this sub-sample of men, was the biomarker interview and testing for level of haemoglobin in blood among children 6-59months of age. The present study focused on this sub-sample of children aged 6-59months on whom blood haemoglobin concentration assessment was made. Additional details about the organization and conduct of the 2018 NDHS are described in the final report^6^

## Key Variables

### Outcome variable

The main outcome (dependent) variable was presence of anaemia, defined as haemoglobin (Hb) level of <11 g/dL^2^

### Independent variables

The major independent/predictor variables considered were categorized into individual (including maternal, paternal and child characteristics), household and contextual/neighbourhood variables. The maternal variables considered included maternal age, maternal education, maternal ethnicity, marital status, religion, maternal autonomy; for the paternal, only paternal education was considered. While the child’s variables included child’s age in months (6-59months), child’s sex, place of delivery, birth weight, birth order, attendance at antenatal care (ANC), uptake of vitamin A supplementation in past 6months, presence or absence of wasting, stunting and underweight, experiencing fever or episode or diarrhoea in past two weeks. The household variables included wealth quintile, household size, number of children under five years, source of energy, source of drinking water and type of material used in house floor. The contextual/neighbourhood variables are place of residence and geopolitical zone. For the detailed description of variables see table 1.

**Table 1.**
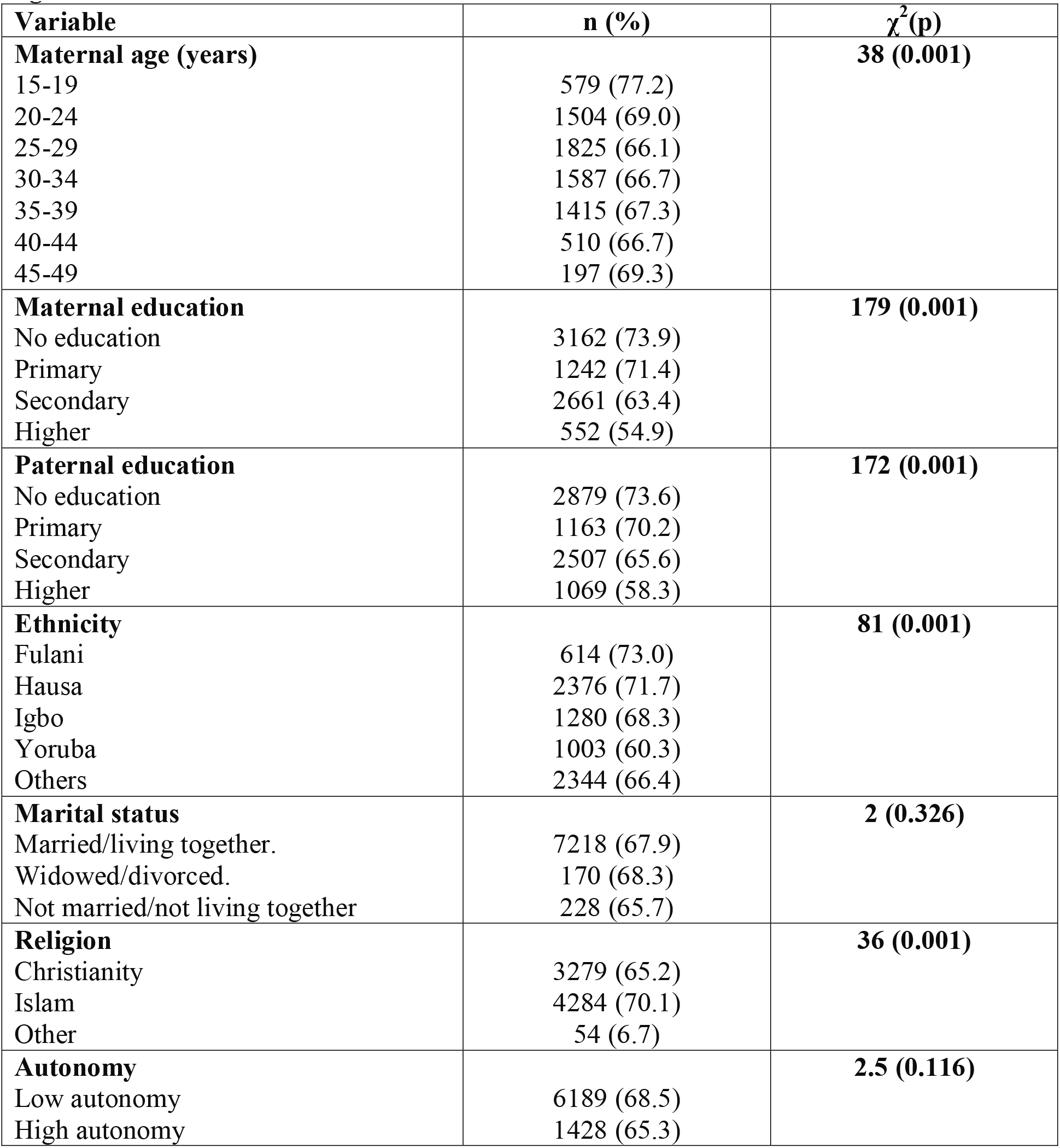
Prevalence of anaemia by background parental sociodemographic characteristics, Nigeria DHS 2018.

| **Variable** | **n (%)** | **$\chi^2$(p)** |
| --- | --- | --- |
| **Maternal age (years)** |  | **38 (0.001)** |
| 15-19 | 579 (77.2) |  |
| 20-24 | 1504 (69.0) |  |
| 25-29 | 1825 (66.1) |  |
| 30-34 | 1587 (66.7) |  |
| 35-39 | 1415 (67.3) |  |
| 40-44 | 510 (66.7) |  |
| 45-49 | 197 (69.3) |  |
| **Maternal education** |  | **179 (0.001)** |
| No education | 3162 (73.9) |  |
| Primary | 1242 (71.4) |  |
| Secondary | 2661 (63.4) |  |
| Higher | 552 (54.9) |  |
| **Paternal education** |  | **172 (0.001)** |
| No education | 2879 (73.6) |  |
| Primary | 1163 (70.2) |  |
| Secondary | 2507 (65.6) |  |
| Higher | 1069 (58.3) |  |
| **Ethnicity** |  | **81 (0.001)** |
| Fulani | 614 (73.0) |  |
| Hausa | 2376 (71.7) |  |
| Igbo | 1280 (68.3) |  |
| Yoruba | 1003 (60.3) |  |
| Others | 2344 (66.4) |  |
| **Marital status** |  | **2 (0.326)** |
| Married/living together. | 7218 (67.9) |  |
| Widowed/divorced. | 170 (68.3) |  |
| Not married/not living together | 228 (65.7) |  |
| **Religion** |  | **36 (0.001)** |
| Christianity | 3279 (65.2) |  |
| Islam | 4284 (70.1) |  |
| Other | 54 (6.7) |  |
| **Autonomy** |  | **2.5 (0.116)** |
| Low autonomy | 6189 (68.5) |  |
| High autonomy | 1428 (65.3) |  |

## Statistical method

### Data preparation

For this study, two data files were utilized: the household members (or Person) recode file (NGPR7AFL) and kids recode file (NGKR7AFL). The NGPR7AFL allows for the estimation of the overall prevalence of childhood anaemia together with some of the selected household variables as outlined in the previous section above. The NGKR7AFL allows for the estimation of other individual and contextual/neighbourhood variables as specified above. Thus, after generating these relevant variables (individuals, household, and contextual/neighbourhood), the two data files were merged into one single file to allow further statistical analyses.

The statistical analysis was conducted in two stages. The first stage was the generation of descriptive statistics. We estimated the overall prevalence of anaemia and cross-tabulate this prevalence with some socio-demographic characteristics to determine if there were associations between them and presence of anaemia among the children. These results are presented in Table 1. The second-stage analysis was the multi-level logistic regression. The data structure is hierarchical with individuals (children, level 1) nested within household that were further nested within context environment/neighbourhood (level 2). This hierarchical nature of the data allowed us to perform the multi-level modelling of the factors associated with anaemia among under-five children in Nigeria. Here, the contextual/neighbourhood variable is the cluster. This ‘cluster’ could equally be assumed to be the ‘community’, ‘residential areas’, or ‘neighbourhood’^9^ and we aimed to investigate whether cluster of residence has any effect on child’s haemoglobin concentration, i.e., if cluster of residence determined presence or absence of anaemia among under-five children in Nigeria.

### Model Development

Multi-level logistic regression model has two components: fixed and random effects parts. This can be simplified as follows.^10^

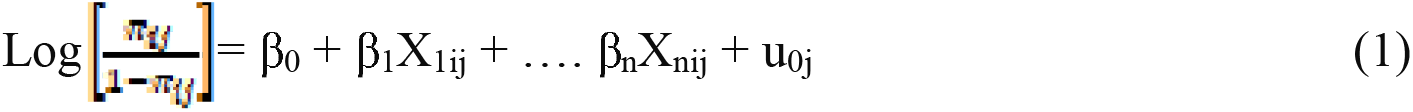

Where π*_ij_*is the proportion of under-five children with anaemia and (*1-*π*ij*) is the proportion without anaemia.

β*_0_* is the intercept coefficient.

β*_1_, …* β*n* are the coefficients of individual and contextual/neighbourhood-level factors *X_1ij_… X_nij_* are independent variables of individuals and contextual/neighbourhood. *u_0j_* are random errors at cluster levels/contextual/neighbourhoods. Equation (1) above can also be simply written as^11^:

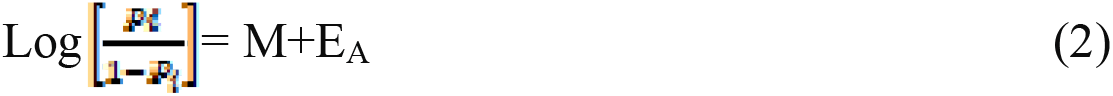

Where *P_i_* is the probability of anaemia in under-five child M=overall mean probability (prevalence) expressed on the logistic scale. E_A_=area (cluster/neighbourhood) level residual. The area level residuals are on the logistic scale and normally distributed with mean 0 and variance V_A_. V_A_=area residual variance expressed on the logistic scale (that is, variance around M). To measure the contribution of cluster/neighbourhood and clustering effects on the prevalence of anaemia among under-five children, we estimated intra-class correlation coefficient (ICC) and the closely related variance partition coefficient (VPC). Thus, there are variations between ‘neighbourhoods’ or ‘clusters’ and there are variations between children within the same ‘clusters’ or ‘neighbourhoods’ and these total variations contributed to the level of anaemia in under-five children as well. Intra-cluster correlation coefficient is a measure of the relatedness of the under-five children in their own cluster/neighbourhood and it measures the share of the total outcome variance which lies at the context/neighbourhood/cluster level, having adjusted for any covariates; an ICC of ‘0’ indicates that individuals within clusters are no more similar to each other than individuals from different clusters (there is no between-cluster/between-neighbourhood variability), while an ICC of ‘1’ indicates that individuals within the same cluster all have identical outcomes (there is no within-cluster variability or the individuals are homogenous.^12^ Intuitively, it is the ratio between cluster variability and total cluster variability (i.e., between-cluster and within-cluster variability). Thus, it is represented by rho (ρ)^12^:

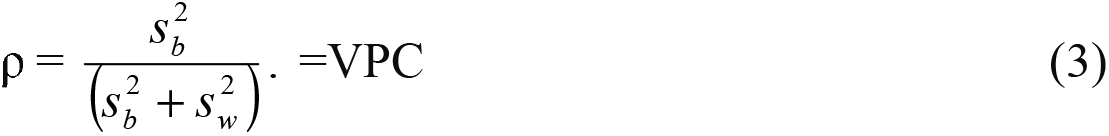

Where 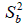 is the variance between clusters, while 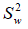 is the variance within clusters. This equation can also be represented as follows:

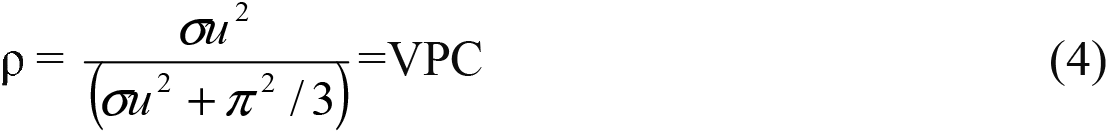

In standard logistic distribution, π^2^/3 = 3.29.^13,14^

To measure the contribution of cluster/neighbourhood effect on prevalence of anaemia among under-five children’s, proportional change in variance (PCV) was used. It is calculated using the ‘empty’ or ‘null’ model as a reference model. It is mathematically computed as:

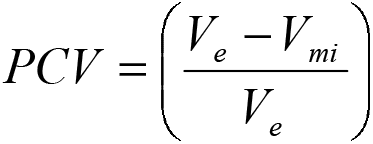

Where, *V*_e_ is the variance in prevalence of anaemia in the empty model and *V*_mi_ is the variance in prevalence of anaemia in model m*_i_* (or the subsequent model). To model the effect of ‘cluster’ or ‘neighbourhood’ variability/characteristics on the occurrence of anaemia among under-five children in Nigeria, we developed multi-level logistic regression analysis (MLRA) in stages. These stages are:

1. **Null or Empty model (Model I):** This is called ‘empty’ model since it does not contain any explanatory/independent variable. It estimates only the average prevalence of anaemia and the neighbourhood and individual differences in anaemia.
2. **Model II:** In this model, only individual characteristics are added to the model; it estimated the effect of individuals (under-five children) on the prevalence of anaemia.
3. **Model III:** This model contains the household-level factors. It thus assessed the effects of household factors as they influenced occurrence of anaemia.
4. **Model IV:** This model has the neighbourhood/contextual characteristics in the model. These contextual characteristics are the place of residence and geopolitical zone.
5. **Model V:** This model has Models I-III added; that is, individual, household, and contextual factors added to determine their combined effects on the prevalence of anaemia. This model was built sequentially by adding one contextual-level variable at a time to avoid potentiality of collinearity with other variables and to see if addition of contextual-level variables improves the model.

All analyses were conducted using Stata version 14 (StataCorp).

## Results

### Descriptive analysis

Data on a weighted sample of 11,223 under-five children in approximately 42,000 households across the 6 geopolitical zones of Nigeria were analysed. The overall prevalence of anaemia was 67.9%. Chi-squared test and cross-tabulation (Table 1 & 2) demonstrated a statistically significant association between the outcome variable (anaemia in under-five children in Nigeria) and most of the predictor variables at the individual, household, and contextual/neighbourhood levels (maternal age, maternal education, paternal education, ethnicity, religion, birth weight, child’s age, birth order, ANC visit, wasting, stunting, underweight, vitamin A supplementation, fever in the past 2 weeks, wealth quintile, household size, number of under-five children, type of sanitation/water source, household energy, floor material, geopolitical zone, and place of residence). The only independent variables found not to be significantly associated with anaemia were marital status (p>0.326), autonomy (p>0.116), sex of the child (p>0.701), and diarrhoea in the past 2 weeks (p>0.365).

Under-five children with anaemia were more likely to be 24-59-month-old, wasted, stunted, underweight, or to have had a recent fever. These children were also more likely to have been delivered at home as either first or fifth child to low-income parents living in homes with earth/sand/dung floors, having low level of education, large household size, and poor water/energy supply and sanitation. Additionally, the children were more likely to have very young mothers (15-19 years old) or perimenopausal mothers (45-49 years old), resident in rural areas in the southwest, northeast, or northwest geopolitical zone of Nigeria.

There was a comparative decreasing prevalence of anaemia with increasing maternal (from 75% to 55%) and paternal educational levels (from 74% to 58%). Similarly, decreasing prevalence of anaemia was observed with increasing number of ANC visits (from 76% to 65%) and increasing wealth quintile (from 78%, 72%, 68%, 67% to 54%).

### Multivariable Multilevel Logistic Regression (MMLR) Analyses

Across all the five multivariable logistic regression models, ten variables were significantly associated with anaemia. These included maternal education, religion, order of birth, age, delivery at health facility, undernutrition states (wasting, stunting, and underweight), household size, and number of under-five children, wealth quintile, floor material, region, and area of residence.

In the final multivariable multilevel logistic regression (MLR) model (Model V, Table 3), the individual, household, and neighbourhood factors significantly associated with increased odds of anaemia among under-five children are: having Muslim parents [OR: 1.33; 95% CI: (1.14-1.55)], a child being big baby at birth [OR: 1.34; 95% CI: (1.04-1.73)], a child being wasted [OR: 1.36; 95% CI: (1.11-1.69)], a child being stunted [OR: 1.32; 95% CI: (1.18-1.48)], a child being underweight [OR:1.29; 95% CI: (1.12-1.49), being a child resident in Southeastern [OR: 1.98; 95% CI: (1.61-2.43)] and Southwestern Nigeria [OR: 2.15; 95% CI: (1.73-2.67)]; and being a child resident in rural area [OR: 1.17; 95% CI: (1.03-1.34)].

**Table 2.** Prevalence of anaemia by child characteristics, Nigeria DHS 2018.

| **Variable** | **n (%)** | **$\chi^2$(p)** |
| --- | --- | --- |
| **Sex of child** |  | **0.15 (0.701)** |
| Male | 3788 (67.4) |  |
| Female | 3829 (68.3) |  |
| **Birthweight** |  | **101 (0.001)** |
| Low birth weight | 88 (54.3) |  |
| Normal | 1550 (60.6) |  |
| Big baby | 344 (67.8) |  |
| **Age of child (months)** |  | **8 (0.016)** |
| 6-23 | 651(64.4) |  |
| 24-59 | 2147 (68.3) |  |
| **Birth order.** |  | **24 (0.001)** |
| First | 2299 (68.0) |  |
| 2<sup>nd</sup> – 4<sup>th</sup> | 3251 (65.5) |  |
| Fifth plus | 2067 (71.9) |  |
| **Place of delivery** |  | **130 (0.001)** |
| Home | 4393 (72.4) |  |
| Health facility | 3225 (62.6) |  |
| **Number ANC visits.** |  | **70 (0.001)** |
| None | 872 (76.0) |  |
| 1-3 visits | 630 (73.2) |  |
| 4+ | 2487 (64.7) |  |
| **Wasting status** |  | **38 (0.001)** |
| Not wasted | 6984 (67.1) |  |
| Wasted | 601 (79.2) |  |
| **Stunting status** |  | **147 (0.001)** |
| Not stunted | 4326 (63.6) |  |
| Stunted | 3225 (74.8) |  |
| **Underweight status** |  | **138 (0.001)** |
| Not underweight. | 5655 (65.0) |  |
| Underweight | 1949 (77.9) |  |
| **Vitamin A supplementation past 6 months** |  | **23 (0.001)** |
| None | 4514 (69.3) |  |
| Yes | 3103 (65.9) |  |
| **Fever past 2 weeks** |  | **7 (0.007)** |
| No fever | 6131 (66.9) |  |
| Yes, last 2 weeks | 1486 (72.0) |  |
| **Diarrhoea past 2 weeks** |  | **0.8 (0.365)** |
| No diarrhoea | 6998 (67.8) |  |
| Yes, last 2 weeks | 619 (69.3) |  |
| **Region** |  | **95 (0.001)** |
| North central | 1044 (65.9) |  |
| North east | 1262 (70.3) |  |
| North west | 2212 (69.9) |  |
| South east | 1036 (69.8) |  |
| South west | 861 (73.2) |  |
| South south | 1203 (59.6) |  |
| **Residence** |  | **133 (0.001)** |
| Urban | 3066 (62.0) |  |
| Rural | 45551 (72.5) |  |
| **Wealth quintile** |  | **271 (0.001)** |
| Poorest | 1612 (77.9) |  |
| Poorer | 1557 (72.1) |  |
| Middle | 1555 (68.3) |  |
| Richer | 1784 (67.4) |  |
| Richest | 1108 (53.6) |  |
| **Household size** |  | **32 (0.001)** |
| 1-5 | 3062 (65.2) |  |
| 6 and more | 4555 (69.8) |  |
| **Number of under-five children** |  | **8 (0.004)** |
| 0-4 | 7271 (67.7) |  |
| 5 and more | 346 (72.1) |  |
| **Type of sanitation/water source** |  | **155 (0.001)** |
| Non-improved | 3754 (74.9) |  |
| Improved | 3863 (62.2) |  |
| **Household energy source** |  | **129 (0.001)** |
| Solid fuels | 6721 (70.1) |  |
| Clean fuels | 719 (51.9) |  |
| Others | 177 (70.0) |  |
| **Floor material** |  | **198 (0.001)** |
| Earth/sand/dung | 2445 (77.0) |  |
| Polished wood | 10 (44.4) |  |
| Tiles | 434 (53.4) |  |
| Cement | 4491 (666.2) |  |
| Carpet | 235 (55.7) |  |
| Others | 2 (45.8) |  |

**Table 2.**
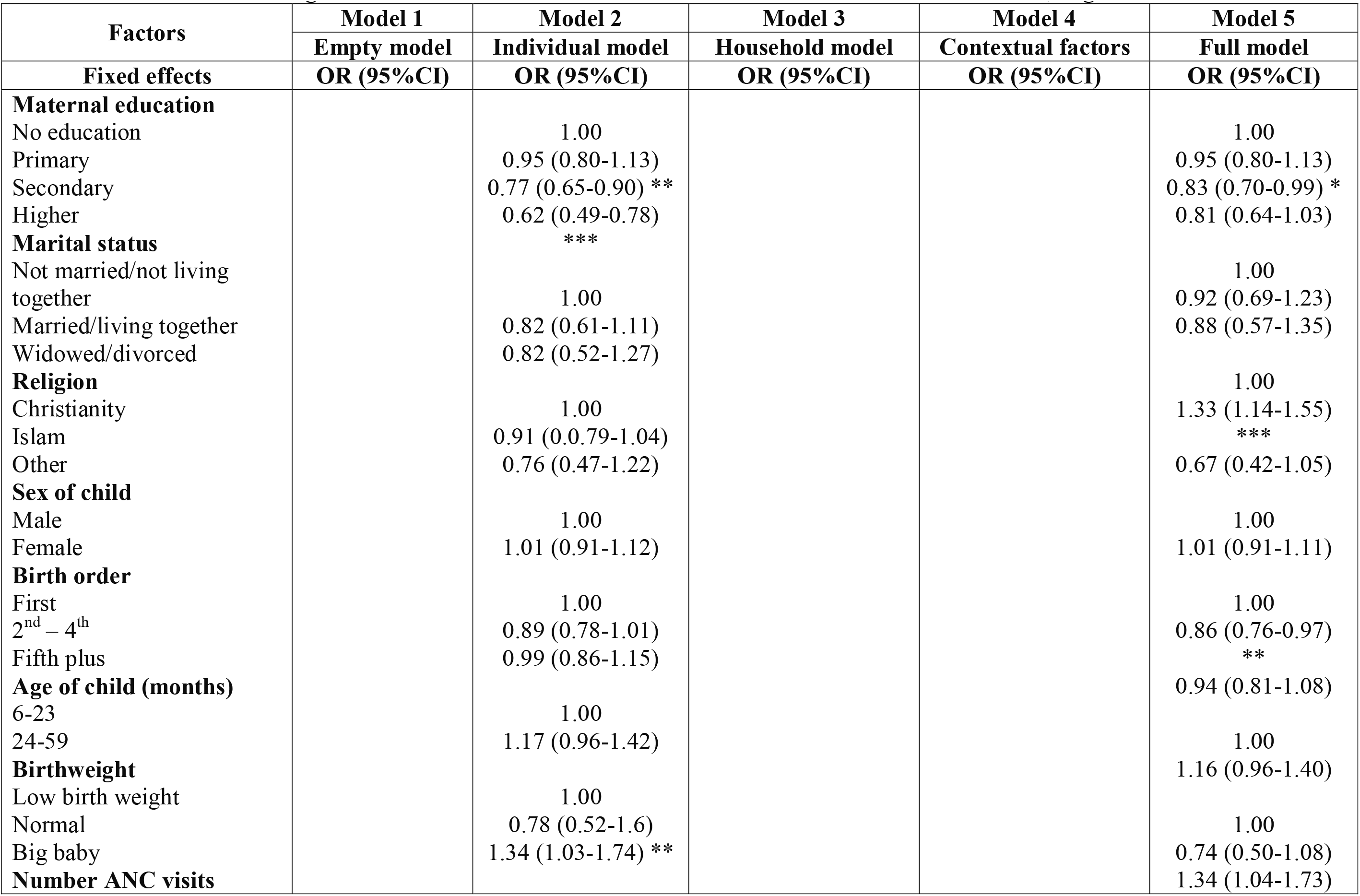
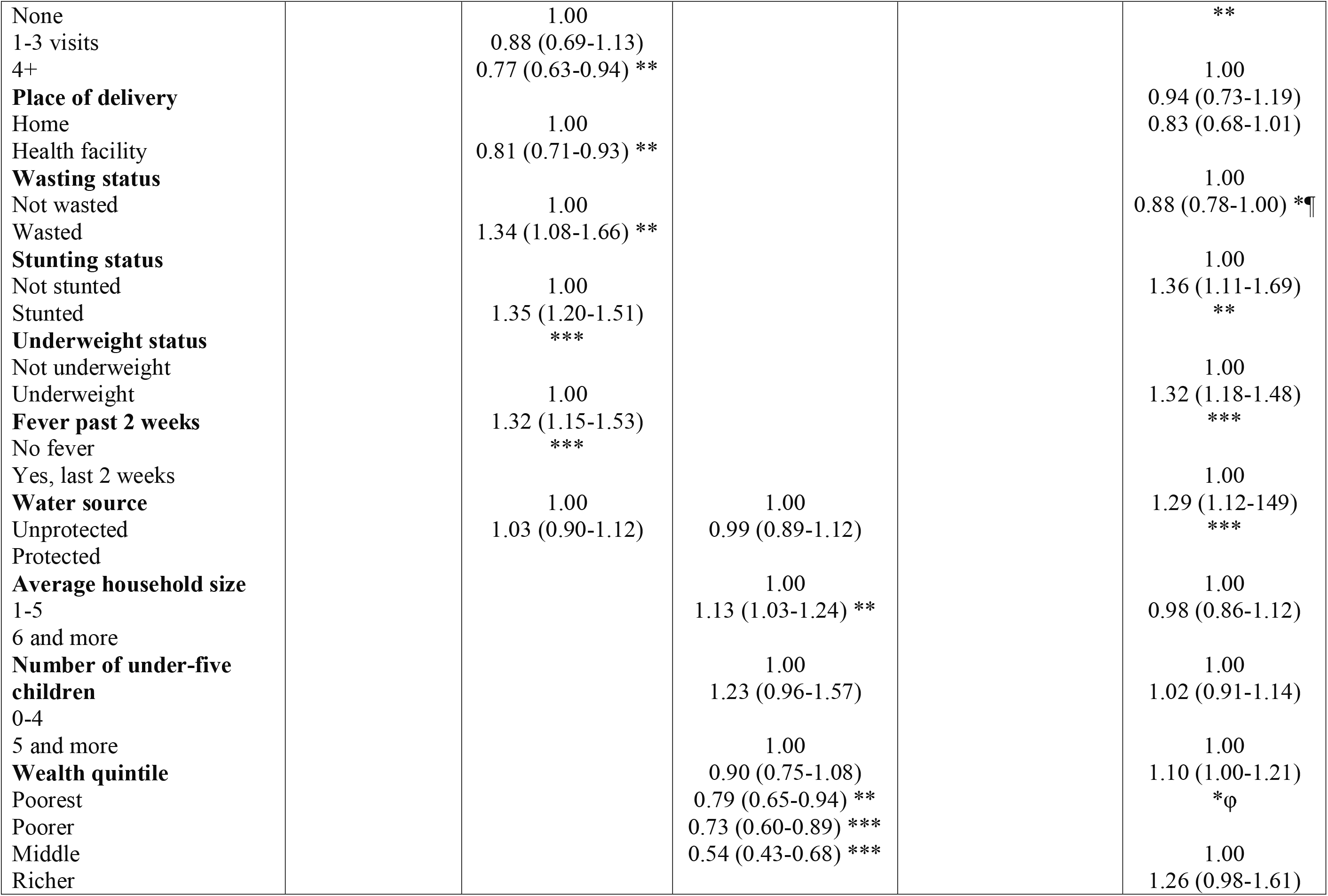
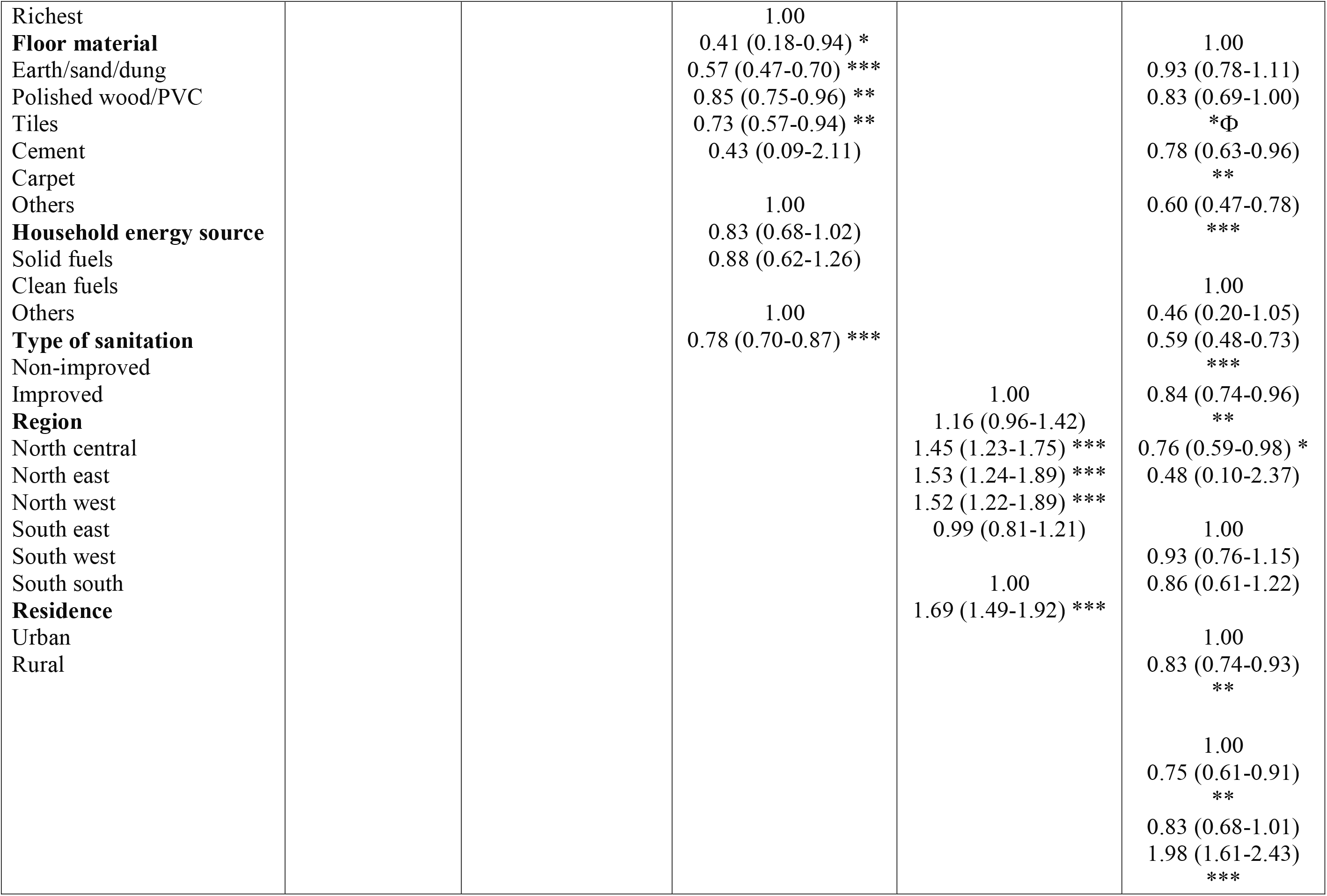
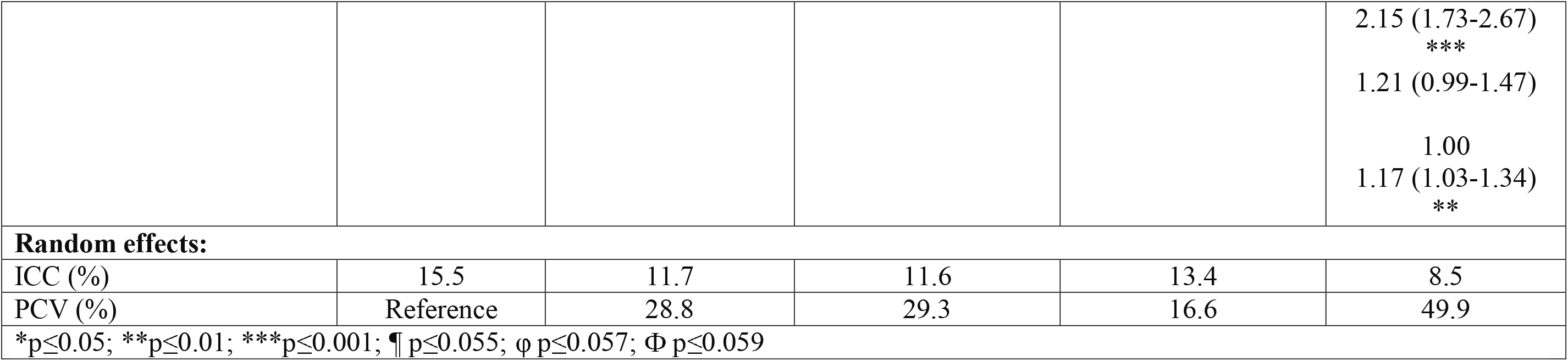
Multilevel regression model of factors associated with anaemia in under-five children, Nigeria DHS 2018.

On the other hand, factors significantly associated with decreased odds of anaemia included are: a mother with secondary education [OR: 0.83; 95% CI: (0.70-0.99)], a child in the second, third, or fourth birth order [OR: 0.86; 95% CI: (0.76-0.97)], belonging to richest households [OR: 0.60; 95% CI: (0.47-0.78)], living in households with tiled floors [OR: 0.59; 95% CI: (0.48-0.73)], living in households with cemented floors [OR: 0.84; 95% CI: (0.74-0.96)], living in households with carpeted floors [OR: 0.76; 95% CI: (0.59-0.98)], and improved sanitation/water source [OR: 0.83; 95% CI: (0.74-0.93)] and living in Northeast [OR: 0.75; 95%CI: (0.61-0.91)].

The odds of developing anaemia in under-five children decreased consistently with increasing levels of maternal education; hence children whose mothers had primary, secondary and higher education had 0.95, 0.77, and 0.62 times less odds for having anaemia compared to those with uneducated mothers. Similarly, odds of anaemia decreased steadily with increasing wealth quintile from middle to richer and to the richest (0.83, 0.78, and 0.60, respectively).

The multilevel logistic regression (MLR) models indicate values of ICC ranging from 8.5% (in Model 5) to 11.7% in Model 2. In Model 1 (or the Null/Empty Model), the ICC is 15.5% showing that average prevalence of anaemia of 15.5% is due to variation/differences in individual, household and contextual (community or neighbourhood) factors. Similarly, in Model 2, the ICC is 11.7% indicating that average prevalence of anaemia due to individual variation is 11.7%; while Model 3 show an ICC of 11.6% and Model 4 show an ICC of 13.4%. The final model (Model 5) shows an ICC of 8.5% indicating that the combination of individual, household and contextual factors determine the average prevalence of anaemia in children in Nigeria.

We estimated the contributions of individual, household, and neighbourhood (cluster) effects on prevalence of anaemia among under-five children using PCV (Proportional Change in Variance). Table 3 shows that these estimates range from 16.6% in Model 4 to 49.9% in Model 5. The combined effects of individual, household and contextual factors accounted for 50% of the determinants of anaemia in children. The quantification of this ‘household, and or ‘neighbourhood’ phenomenon has some implications in terms of control and prevention.

## Discussion

This study employed a nationally representative data and found a 67.9% overall prevalence of anaemia among the weighted sample of 11,223 under-five children in Nigeria. This is unacceptably high and indicates anaemia as a severe public health problem in this age category as classified by the WHO.^2^ Other studies across Africa reveal similarly high prevalence ^15–17^, we observe an average regional prevalence of 59.9% for sub-Saharan African with children under 10 years of age suffering the highest burden.^4,18^ The pervasive level of anaemia in these young children prevents them from attaining optimal levels of cognitive and motor achievements, leading to population of adults with markedly reduced productivity and earning potentials.^19^

The results of this study demonstrate an association between anaemia in under-five children and most of the child, maternal and community factors. The children with anaemia were older, had undernutrition, suffered a recent illness (likely upper respiratory infection or malaria fever), were children of teenage mothers or women much older who had delivered them at home. Anaemia prevalence was found among children from larger, poor households living in rural areas within poor environments in Nigeria. On the other hand, anaemia levels were lower among children whose parents had higher levels of education, whose mothers had higher numbers of ANC contacts and were from households of higher socioeconomic status. Similarly, other studies found that sociodemographic factors, socioeconomic factors, child nutrition and growth, parental factors and family structure, environmental factors as well as recent illnesses all contribute to anaemia among young children.^4,5,15,16^

In the present study we found that child’s birth order, undernutrition, place of delivery, maternal education, household size, wealth quintile, housing floor material, religion, place, and region of residence all remained significantly associated with anaemia in children across all the five multivariable logistic regression models. While in the final model, factors that were associated with increased odds of anaemia among these children were having Muslim parents, being a big baby at birth, being under nourished, living in a rural area or resident in Southeastern or Southwestern region of Nigeria.

Anaemia is an important outcome indicator of poor nutrition and health with young children and pregnant women most vulnerable sub-population group. Adolescent and very young women who may not be physiologically ready to provide the nutritional needs of their children due to lack of adequate micronutrient stores, especially iron, go on to have first babies with anaemia, on the other hand, children born to older or perimenopausal women who likely might have had many or too frequent and poorly spaced pregnancies are more likely to be anaemic. Studies reveal that women have a higher burden of anaemia than men of same age due to regular blood losses during menstruation and childbirth that result in depleted iron stores.^3,17,20^ Giving the contribution of maternal anaemia to child anaemia status, reducing the prevalence of anaemia among adult women and adolescent girls will greatly add towards reduction of prevalence of anaemia in children.^21^

The present study revealed that children with stunting, wasting or underweight were more likely to have anaemia. Infants and young children, particularly during the first two years of life experience rapid growth and development which becomes compromised in the presence of anaemia. Unfortunately, typical complementary foods are frequently monotonous and low in iron content. In addition, childhood morbidities such as diarrheal diseases, malaria fever and upper respiratory infections can cause anaemia through impaired nutrient absorption and metabolism or increased nutrient losses.^22^ Thus, in the presence of a compromised growth of the child, the synergistic association between nutrient deficiency and anaemia in young children greatly flourishes.^23^

This study found a significant association between maternal education and anaemia in young children. Maternal educational attainment has been shown to influence childcare and feeding practices.^3,24^ Children were less likely to have anaemia with increasing levels of maternal education, this is likely due to the finding of a positive relationship between maternal education, knowledge of childcare and improved child feeding.^4,5,7,8,18,24^ The present study also found that household size, which is linked to the wealth quintile, was an important contributor to a child having anaemia. This is similar to another study in Ethiopia where larger households had limited resources, especially in presence of many children under five years, with resultant increased food insecurity and childhood anaemia.^24^

The final model of this study showed prevalence of anaemia in babies born big. This is different from another study which showed that likelihood of being anaemic to be higher among low birth weight (LBW) children.^5,8,24^ This may be explained by the fact that despite being big at birth, subsequent nutritional inadequacies over time and likely repeated infections may deplete the stores and lead to development of anaemia. This final model also revealed that children of parents who were Muslims, who lived in rural areas or who were residents of Southeast and Southwest Nigeria were more likely to have anaemia. These could be explained by several factors that include practices that are peculiar to religious groups, for instance, large household sizes may be more common among Muslims, also, a large proportion of the Muslims in Nigeria are Hausa/Fulani and may consume milk which may reduce the bioavailability of iron as shown in a study from Tanzania.^25^ Other explanations are the significant differences in the infant and young child feeding practices, culture, and community norms and not least importantly the socioeconomic status which informs the living standards of the people in these areas and regions of residence. Similar findings were seen in Ghana and Ethiopia.^5,26^

Anaemia was much less likely to be found among children belonging to the richest households, where the homes had tiled, cemented, or carpeted floors, and lived in homes with an improved water source and sanitation. The significant association between poverty and anaemia is explained by decreased household food security, inadequate maternal and childcare, decreased access to health services while living in poor environments that increases risk of infections, further causing nutritional compromise.

The multilevel logistic regression models used in this study have indicated that the average prevalence of anaemia in children in Nigeria is due to variations in individual, household, and contextual factors. The effects of these factors, as shown by estimates of proportional change in variance (PCV) ranged from 16.6% to 49.9%, thus this gives a combined account of about 50% of the determinants of anaemia in children in Nigeria. The findings of this study can be quantified and applied into practice, with great implication for prevention and control. A reduction in the prevalence of anaemia among children in Nigeria will lower the economic burden, this in turn will alleviate poverty and the improve productivity of the population. A multidimensional approach towards reduction of anaemia in young children may employ health promotion and behaviour change interventions, specific disease prevention and control as well as agricultural and nutritional strategies.

The findings from this study reveal that individual, household and contextual factors are important determinants of anaemia in children in Nigeria. It is imperative to quantify these factors and identify the multidimensional strategies to address them; this will help inform policy decisions towards reduction of effects of anaemia on child growth and development.

## KEY MESSAGES

Anaemia among children is an issue of significant public health concern with higher burden in low– and middle-income countries. This study identified several individual, household, and contextual determinants of anaemia among children under five years. These findings will aid evidence-based child health policy decisions towards reduction of effects of anaemia on child growth and development.

## STRENGTHS AND LIMITATIONS OF THIS STUDY

The study used advanced statistical techniques to validate the association between several individual, household, and contextual determinants of anaemia among children under five years. Being a household survey; recall bias may be a limitation. Further, the survey might have missed other factors associated with anaemia in children. Also, determining cause-effect relationship may not be possible with a cross-sectional study. Finally, there is need for further studies to aimed at further unravelling the very complex interrelationships between and among these individual, household, and contextual factors of anaemia in young children in Nigeria.

## Data Availability

All data used in preparing this manuscript are available online at https://www.dhsprogram.com/data/dataset/Nigeria_Standard-DHS_2018.cfm?flag=0

